# Efficacy of behavioural activation in treating prenatal depression: a systematic review and meta-analysis

**DOI:** 10.1101/2025.07.09.25331226

**Authors:** Engida Yisma, Kuda Muyambi, Shwikar Othman, Sandra Walsh, Dan Bressington, Mary Steen, Richard Gray, Jennifer Fereday, Zohra Lassi, Martin Jones

## Abstract

**Background:** Prenatal depression is a significant and increasingly recognised public health issue, with symptoms often intensifying as pregnancy progresses. Behavioural activation is an established treatment for depression; however, its effectiveness in treating depression during the prenatal period has not been systematically evaluated.

**Objective:** To compare the effectiveness of behavioural activation against any comparators in reducing depressive symptoms in pregnant women.

**Methods:** We searched six primary databases (CINAHL, Cochrane Library, Ovid Embase, Ovid Emcare, Ovid MEDLINE, and Ovid PsycINFO) for randomised controlled trials (RCTs) comparing behavioural activation with comparators for depression in pregnant women. We also searched for major international clinical trial registries and manually reviewed reference lists of included studies. Risk of bias was assessed using the Cochrane Collaboration’s risk-of-bias 2 tool. We calculated a random-effects, inverse-variance weighting meta-analysis to examine the association between behavioural activation and depressive symptoms during the prenatal period.

**Results:** Of the 4,129 records identified, four RCTs from three countries comparing behavioural activation with comparators for the treatment of depression in pregnant women met the inclusion criteria. One study was rated as low risk of bias. Meta-analysis showed that behavioural activation was associated with a small reduction in depressive symptoms among pregnant women diagnosed with depression, compared to treatment as usual (standardised mean difference = −0.33; 95% CI, −0.57 to −0.10), with high heterogeneity across studies (I² = 82.8%). Two of the four included studies reported on adverse events; however, none reported any harm resulting from participation in the treatment trials.

**Conclusions:** This systematic review indicated that behavioural activation may be more effective than treatment as usual in reducing depressive symptoms during the prenatal period. However, the findings should be interpreted with caution due to limitations in the quality of the available evidence.

## Introduction

Depression is common in women during the perinatal period, with the prevalence rates tending to progressively increase between the first (7.4%) and second trimesters of pregnancy (12.8%) [1]. In a retrospective cohort study of 17, 564 mothers of infants born in public health facilities in New South Wales (NSW), Australia, a prevalence rate of 7% of antenatal depressive symptoms was identified [2]. The same study identified associations between prenatal and postnatal depressive symptoms [2]. In a systematic review of 16 studies that involved 35,419 women, the authors reported that the average rates of antenatal and postnatal depression across these studies were 17% and 13%, respectively [3]. The same study identified associations between prenatal and postnatal depression with 9% of women who experienced antenatal depression going on to have postnatal depression, while approximately 50% of those with postnatal depression had previously experienced antenatal depression [3]. Another meta-analysis of 101 studies reported the pooled prevalence of perinatal depression at 11.9% [4]. The foregoing evidence from past studies demonstrates that antenatal and postnatal depression impose a substantial burden on women that warrants attention.

Depressive symptoms during the pre-and postnatal periods usually present as emotional (low mood, loss of confidence), cognitive (low sense of self-worth, feeling guilty), behavioural (loss of interest in activity), social (withdrawal from family or friends), and physical (poor appetite, loss of sleep) changes that can cause complications across the perinatal period [5]. The symptoms commonly comprise feelings of sadness, a sense of worthlessness or guilt, loss of interest or pleasure in doing things routinely enjoyed in everyday life, fatigue, poor sleep, and a change in appetite [6]. Symptoms specific to depression during the antenatal period include negative thoughts concerning motherhood and the new baby, with women frequently displaying self-harm behaviour and suicidal ideation [6].

In pregnant women, depression is associated with low utilisation of antenatal health services, frequent hospitalisations, foetal growth retardation, preterm delivery, and low birth weight [7]. Postnatally, depression is known to cause difficulties in undertaking the parenting role with negative consequences for the behavioural development of the offspring [8, 9]. Some studies suggest that early childhood psychological challenges arising from maternal prenatal depression can result in psychiatric disorders, poor academic performance, and social functioning in later adolescent and adult life [10, 11].

Treatment interventions are needed to ameliorate suffering for women experiencing depression or depressive symptoms during the perinatal period. Anti-depressant medication and psychotherapy as monotherapy or in combination are often recommended for treating perinatal depression in women [12]. However, due to reluctance to take antidepressants during breastfeeding based on concerns about potential cognitive difficulties in childhood and maternal side effects (feelings of agitation or anxiety, digestive issues, and headaches), antidepressant medication is less commonly used [13, 14]. Of the available psychotherapies, cognitive behavioural therapy (CBT) is one of the recommended treatments for perinatal depression [15, 16]. CBT involves techniques and strategies to enable people affected by depression to change unhelpful patterns of thinking and behaviour, use problem-solving skills to cope with difficult situations, and develop greater confidence in their abilities [17]. CBT interventions can be delivered in face-to-face one-on-one, group- or internet-based formats [15, 18]. While CBT is effective in treating perinatal depression, it is generally delivered by mental health specialists such as psychologists [18]. This makes CBT expensive to deliver and less accessible in communities that experience shortages and insufficient distribution of the psychologist workforce and other mental health clinicians.

There is strong evidence that behavioural activation, a derivative of CBT, is effective in the prevention and treatment of depression in adults [19, 20]. Evidence also suggests that behavioural activation is equally efficacious as CBT when used as a stand-alone treatment for depression [20]. It is simple and less expensive to deliver by non-specialists [20, 21].

Behavioural activation is based on Martell’s model which emphasises behaviour rather than cognitive change [22]. It is based on the belief that the more people become depressed, the less pleasant experiences they will engage in [23]. Conversely, the fewer pleasurable experiences people have, the more depressed they become [23, 24]. Behavioural activation targets low motivation and focuses on increasing engagement in pleasant and valued social and physical activity to improve mood and disrupt detrimental avoidance [23]. This is achieved by identifying and addressing challenges to the identified valued activities through the development and implementation of a structured behavioural plan aimed to increase motivation and encourage the achievement of identified goals [23]. We speculated that behavioural activation may be a suitable alternative to CBT in treating prenatal depression or depressive symptoms.

### Review objectives and questions

To our knowledge, no studies have reviewed the effectiveness of behavioural activation in treating depression or depressive symptoms during the prenatal period. This systematic review and meta-analysis aimed to summarise the evidence about the effectiveness of behavioural activation compared with usual care in treating prenatal depression. We defined treatment as any intervention initiated prenatally among women who have been diagnosed with depressive symptoms using a self-report or the use of clinician-/researcher-administered clinical assessment scales. The review question was:

*Primary research questions*: Is Behavioural Activation effective in reducing subthreshold depression or major depressive symptoms during women’s prenatal period?

## Methods

The protocol for this systematic review was prospectively registered with the Open Science Framework [25]. We followed The Preferred Reporting Items for Systematic Reviews and Meta-Analyses (PRISMA) guidelines during the preparation and reporting of this review [26].

Accordingly, we developed a PICO (population-intervention-comparison-outcome) mnemonic to describe the key elements of the review.

### Population

Reproductive women diagnosed with subthreshold depression or major depressive symptoms during their prenatal period comprised the population for this review. Participants had to be women aged 18 years or over and of any ethnicity. Subthreshold depression or major depressive symptoms were determined by self-report or the use of clinician-/researcher-administered clinical assessment scales. There was no restriction in terms of sample size.

### Intervention

We included studies that focused on behavioural activation. Studies were included if the treatment was explicitly identified as behavioural activation or its core elements of mood monitoring, and activity scheduling were described as the primary focus of the intervention. The behavioural activation treatment could include face-to-face, online, or group delivery by health practitioners at any level trained in the use of the psychotherapy approach or via self-help. We defined a health practitioner as anyone trained in behavioural activation and registered or approved to provide health care services such as psychologists, psychotherapist, nurse, non-specialist therapists, and community workers. We excluded therapies such as CBT or problem-solving therapy. Studies were ineligible if (a) the intervention involved an experimental pharmacological treatment or component and (b) behavioural activation occurred outside the prenatal period.

### Comparator intervention

Comparator interventions included a waiting list for treatment, usual care, or no treatment.

### Outcome

The main outcome was a change in depressive symptoms as measured by scores from validated self/clinician/researcher-administered clinical rating scales.

### Types of studies

In this review, we included all treatment trials including randomised, cluster-randomised, or non-randomised clinical trials, pilot studies, and feasibility studies. Only studies involving primary research were considered. Studies were included regardless of reported outcomes. We excluded from this review case studies, opinion papers, and systematic and scoping reviews. Systematic and scoping reviews were excluded because we considered them secondary studies [27].

### Clinical setting

We included studies in which the intervention was provided in a hospital, primary health care, or equivalent clinical setting.

### Search strategy

The academic librarian at the University of South Australia assisted with the development of the search strategy. The MEDLINE search string is presented in **Supplementary File** I. Only studies published in English were included in this review to avoid challenges with translation. No date limits were applied.

### Data sources

We searched six primary databases (CINAHL, Cochrane Library, Ovid Embase, Ovid Emcare, Ovid MEDLINE, and Ovid PsycINFO). We also searched the secondary database, Google Advanced (first ten pages). We perused the international trials registries including the WHO International Clinical Trials Registry (https://trialsearch.who.int/), the Australian New Zealand Clinical Trials Registry (http://www.anzctr.org.au/), and ClinicalTrials.gov.trial registries (http://www.ClinicalTrials.gov and the International Standard Randomised Controlled Trial Number Registry (https://www.isrctn.com). The reference lists of retrieved articles were also searched for papers not identified by the database searches. All searches were conducted from May 2022 until October 2022.

### Study selection

We exported the results of the database searches to Endnote reference management software [28] and later uploaded the information to Covidence web-based review management tool [29]. The review management software automatically removed all duplicates. In Confidence, we undertook and completed the title and abstracts, and full-text screening. MJ, SO, EY, and KM independently completed all screenings with the inclusion criteria. Conflicts were discussed by the four-member team until a consensus was reached.

### Data collection and extraction

MJ, SO, SW, EY, and KM independently extracted data from the included studies. These reviewers discussed disagreements amongst themselves until a consensus was reached. We used a data extraction sheet adapted from a template used in past studies by the authors [30]. We extracted study characteristics including study population, setting, recruitment method, treatment, outcome measures, randomisation, sample size calculation, measurements, and effect size.

### Data synthesis and analysis

We performed a narrative synthesis of the findings from the included studies. A meta-analysis was conducted to provide a consolidated estimate of the effect of behavioural activation versus treatment as usual (TAU) on treating prenatal depressive symptoms in women. We synthesised information regarding the reduction in the level of prenatal depressive symptoms following behavioural activation versus treatment as usual by undertaking a random effects meta-analysis using inverse variance weighting. We presented the pooled effect size (standardised mean difference (SMD) with a 95% confidence interval (CI)) and used a forest plot. We used the I^2^ to measure statistical heterogeneity and the possible sources of heterogeneity were explored using posthoc subgroup analyses based on the following subgroups: (a) study design—individually randomised clinical trials, cluster randomised clinical trials, (b) type of outcome measure—PHQ 9 and Hamilton Depression Rating scale, and (c) how the treatment is defined—behavioural activation and variation of behavioural activation. All meta-analyses were conducted using STATA/SE version 17.0 software [31].

### Critical appraisal

The Cochrane risk-of-bias tool Version 2 for randomised trials (RoB 2) was used to assess the risk of bias in the included studies [32, 33]. MJ and EY individually performed the risk of bias assessment and later reconciled their disagreements.

### Ethical approval

This systematic review relied on previously published material and did not require ethical approval.

## Results

Electronic database and clinical trial registry searches identified 4024 published studies, while Google Scholar searches identified a further 100 articles. Five publications were identified using manual searches. We removed 1066 duplicates electronically using Covidence review management software. Title, abstract, and full-text screening accounted for 3059 studies being excluded from the review. Four studies met the inclusion criteria for this systematic review. The study selection process is graphically presented in a PRISMA Flow Chart (**Figure 1**).

**Figure 1.**
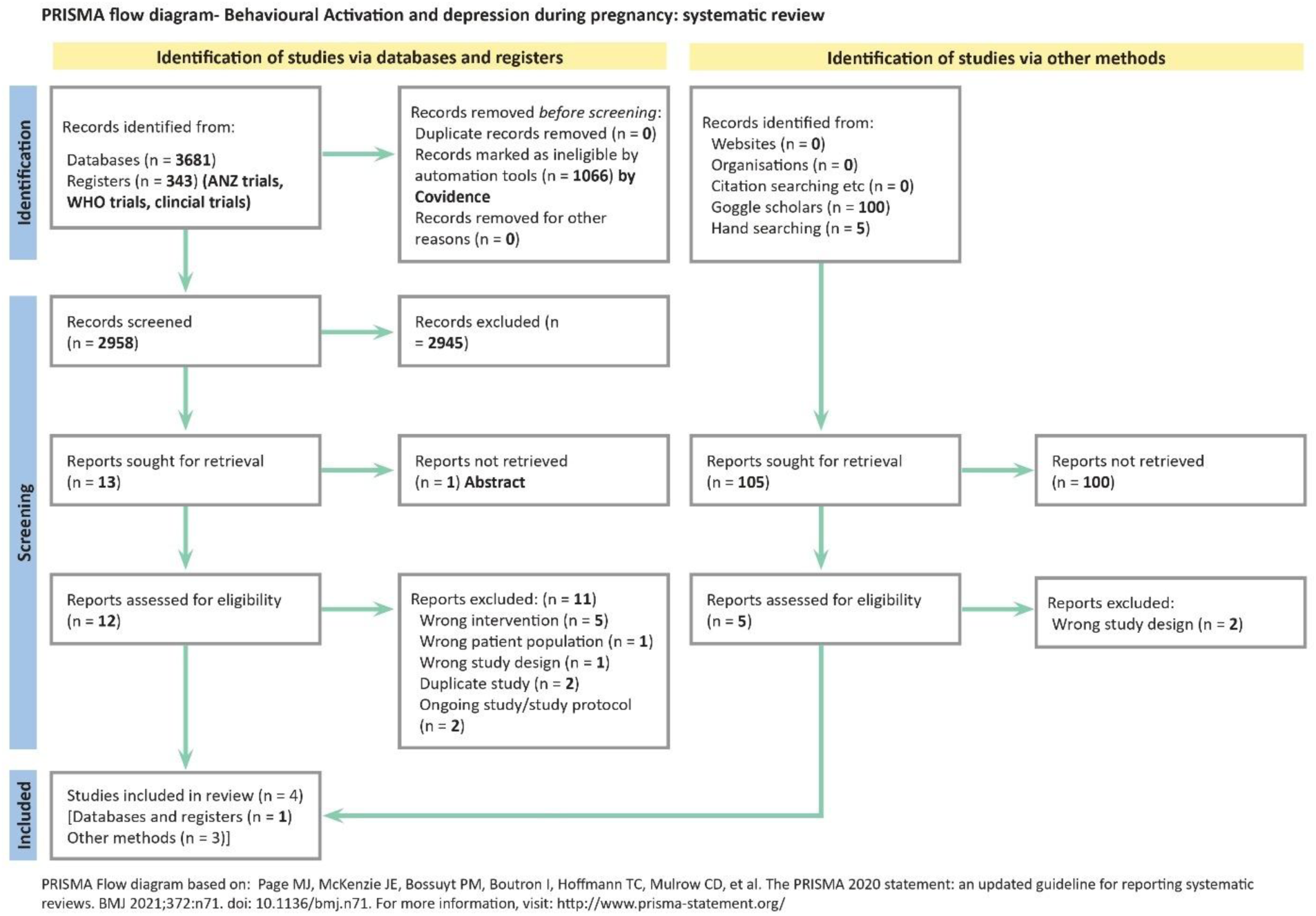
PRISMA Flow chart.

### Description of the included studies

A detailed summary of the four included studies is presented in **Supplementary Appendix 2**. Rahman et al. (2008) examined the effectiveness of the Thinking Healthy Program (THP), a psychological intervention recommended for the treatment of prenatal depression in women during pregnancy [34]. The study, prospectively registered as a trial (ISRCTN65316374), was included because it met our criteria for behavioural activation (BA) since the primary focus was on mood monitoring and activity scheduling [34]. Dimidjian et al. (2017) tested the effectiveness of BA for depression compared with treatment as usual among pregnant women [35]. The trial was prospectively registered at clinicaltrials.gov (NCT01401231). Sikander et al. (2019) assessed the effectiveness and cost-effectiveness of the Thinking Healthy Peer-delivered Program (THPP), a modified THP intervention in treating prenatal depression in women aged 18 years or older [36]. The trial was prospectively registered as number NCT02111915 on ClinicalTrials.gov. Similarly, Fuhr et al. (2019) examined the effectiveness of the Thinking Healthy Peer-delivered Programme (THPP) in the treatment of perinatal depression in pregnant women aged 18 years or above [37]. The trial was prospectively registered with ClinicalTrials.gov, reference NCT02104232.

### Study setting

Two studies were conducted in Pakistan [34, 36], one in India [37], and one in the United States of America (USA) [35].

### Type of study design

All included studies were randomised controlled clinical trials. Three studies adopted a cluster randomised controlled trial design [34–36]. One study was a single-blind, individually randomised controlled trial [37].

### Participants

The four included studies recruited 972 pregnant married women (see **Table 1**) with an average age of 26.71 years (range 25–28.5 years) and a diagnosis of depression. The Rahman et al. study recruited married women aged 16–45 years, with depressive symptoms and in their third trimester of pregnancy [34] whilst Fuhr et al. recruited depressed women aged 18 years or older in their second or third trimester [37]. Sikander et al. recruited women with depressive symptoms aged 18 years or above in their third trimester of pregnancy [36]. In the study by Dimidjian et al., the pregnant women were aged 18 years or above with a diagnosis of depression [35].

**Table 1.** Participant characteristics.

| Participant characteristics | Included studies |  |  |  |  |
| --- | --- | --- | --- | --- | --- |
|  | Sikander et al.<br>N=283 | Fuhr et al.<br>N=140 | Rahman et al.<br>N=463 | Dimidjian et al.<br>N=86 | Total<br>N=972 |
| Age (average) | 26.8 years | 25.04 years | 26.5 years | 28.5 years | 26.71years |
| Married/cohabiting | 283 (100%) | 139 (99%) | 463 (100%) | 62 (72%) | 947 (97%) |
| Full/part-time employment | 20 (7%) | 21 (15%) | Not stated | 61 (71%) | 103 (10%) |
| Highest educational qualification |  |  |  |  |  |
| No formal education | 52 (18%) | 21 (15%) | 184 (40%) | Not stated | 257 (26%) |
| Primary education | 68 (24%) | 55 (39%) | 147 (32%) | 4 (4.65%) | 274 (28%) |
| Secondary education | 145 (51%) | 57 (40%) | 116 (25%) | 19 (22.09%) | 337 (35%) |
| University or higher education | 18 (6%) | 7 (5%) | 16 (3%) | 63 (73%) | 104 (11%) |
| Primiparous | 52 (18%) | 61 (44%) | Not stated | 36 (42%) | 149 (15%) |
| Multiparous | 231 (82%) | 79 (56%) | Not stated | Not stated | 310 (32%) |

### Outcomes and outcome measures

In three studies, the primary outcome was improvement in the severity of depressive symptoms [35–37]. In the Rahman et al. study, improved depressive symptoms comprised the secondary outcome with the primary outcomes being better infant weight and height [34]. The measure for the outcome of interest was the Patient Health Questionnaire-9 (PHQ-9) [35–37] except in the Rahman et al. study. Rahman et al. used the Hamilton Depression Rating Scale to measure depressive symptoms [38] with the clinician-administered Diagnostic and Statistical Manual of Mental Disorders (DSM-IV) being used for initial assessments during recruitment [34]. The Patient Health Questionnaire-9 (PHQ-9) scale has prior evidence of validity and reliability [39, 40]. Other relevant scales used by Dimidjian et al. included the Behavioural Activation for Depression Scale-Short Form (BADS-SF), to measure when and how clients engage in rewarding activities during behavioural activation treatment [41], and the self-report Generalised Anxiety Disorder 7-item Questionnaire (GAD-7) for measuring general anxiety symptoms [35, 42]. Both tools have prior evidence of validity and reliability in the study populations [41, 43, 44]. For all included studies, only the English versions of the scales were used [34, 35, 37, 45].

### Intervention

The Dimidjian et al. study used behavioural activation as the intervention delivered by eight providers: four had a nursing degree (nurse midwife, nurse practitioner), three had a master’s degree in behavioural health, and one was a registered occupational therapist. The providers completed behavioural activation training that included two days of in-person workshops and self-paced reading followed by ongoing weekly group telephone supervision (90 minutes) and individual supervision (30 minutes). Behavioural activation training and supervision were provided by two clinical psychologists/researchers. The behavioural activation intervention was guided by a ten-session behavioural activation treatment manual and delivered by telephone, in an obstetric clinic or in participants’ homes. The main behavioural activation strategies included self-monitoring, structuring, and scheduling of activities, problem-solving skills, and enhanced social support. The use of interpersonal communication skills was emphasised. Sessions were structured and incorporated between-session homework. Treatment fidelity was measured using the Quality of Behavioural Activation Scale (QBAS), a 14-item scale to assess the ability to implement behavioural activation [46]. Participants were required to submit audio recordings of role-play to trained research assistants acting as patients at baseline, post-training, and nine-month follow-up. The audio recordings were later assessed by behavioural activation experts. The intervention participants were followed up for assessment at five and ten weeks post-randomisation and three months postpartum [35].

Rahman et al., Fuhr et al., and Sikander et al. adopted various versions of the Thinking Healthy Program (THP), a psychosocial intervention for perinatal depression. THP was initially conceived based on CBT that incorporated elements of BA. Subsequently, the cognitive element was dropped in preference for behavioural activation due to the complexity of CBT when delivered by lay providers [45]. It is worth noting that the Thinking Healthy Programme (THP) is recommended by the World Health Organization (WHO) and incorporated into its Mental Health Gap Action Programme (mhGAP) as the first-line management of perinatal depression in primary and secondary care settings [45, 47, 48].

Rahman et al. delivered the original THP intervention. The intervention was guided by a written protocol that consisted of weekly sessions held over four weeks commencing in the last month of pregnancy. Subsequently, three sessions were provided in the first postnatal month, and nine one-monthly sessions thereafter. The intervention was delivered by trained village-based primary health workers and emphasised CBT techniques of active listening, family involvement, guided discovery, and homework. The health workers received monthly supervision and monitoring from the researchers [34].

In the Sikander et al. study, the THP was adapted to the Thinking Healthy Peer-delivered Program (THPP) to cater for delivery by volunteer peers working in close collaboration with community healthcare workers that participated in tiered group training and supervision provided by trainers who were not mental health specialists [45]. THPP intervention used psychological strategies including behavioural activation; narratives and images to mitigate unhelpful behaviour and to encourage alternative helpful ones. The participants attended fourteen sessions (ten individual and four group sessions), each lasting 30–45 minutes. The sessions were provided from the third trimester of pregnancy to six months postpartum. The frequency, intensity, and content of the sessions were increased during pregnancy until the third month after childbirth. Ten of the fourteen sessions were delivered during pregnancy and in the first three months after childbirth. The remaining four sessions were delivered between four to six months post-partum [36].

Like the Sikander et al. study, Fuhr et al. adopted the THPP intervention focusing on behavioural activation [37]. The THPP was delivered through six to fourteen individual sessions in four phases over seven to twelve months. The four phases were: (a) prenatal phase (one to six sessions), (b) early infancy (one to four sessions), (c) middle infancy (two sessions; and (d) late infancy, intervention delivered between five to six months after childbirth in two sessions.

Phases (b), (c), and (d) were consistent with the study’s primary focus on infant weight and height. Each session, delivered by laywomen with no formal mental health training, lasted between 30 and 45 minutes. Participation in a minimum of six sessions, with at least one session in each of the four phases, represented treatment completion [37].

### Comparators

In the study by Dimidjian et al., treatment as usual (TAU) consisted of routine care [35], whilst the Rahman et al., Fuhr et al., and Sikander et al. studies involved what was termed ‘enhanced usual care’ involving routine care and testing for depression as the comparators [34, 36, 37]. Reportedly, enhanced usual care involved the provision of usual care by a health practitioner, testing for depression, and making referrals to appropriate services for those testing positive [34, 36, 37].

### Outcomes

The primary outcome of the Rahman et al. study was improved depressive symptoms [34]. At the end of the study, the participants in the intervention group attained lower depression scores at the two time points compared to those in the control group [34] (**Table 2**).

**Table 2.**
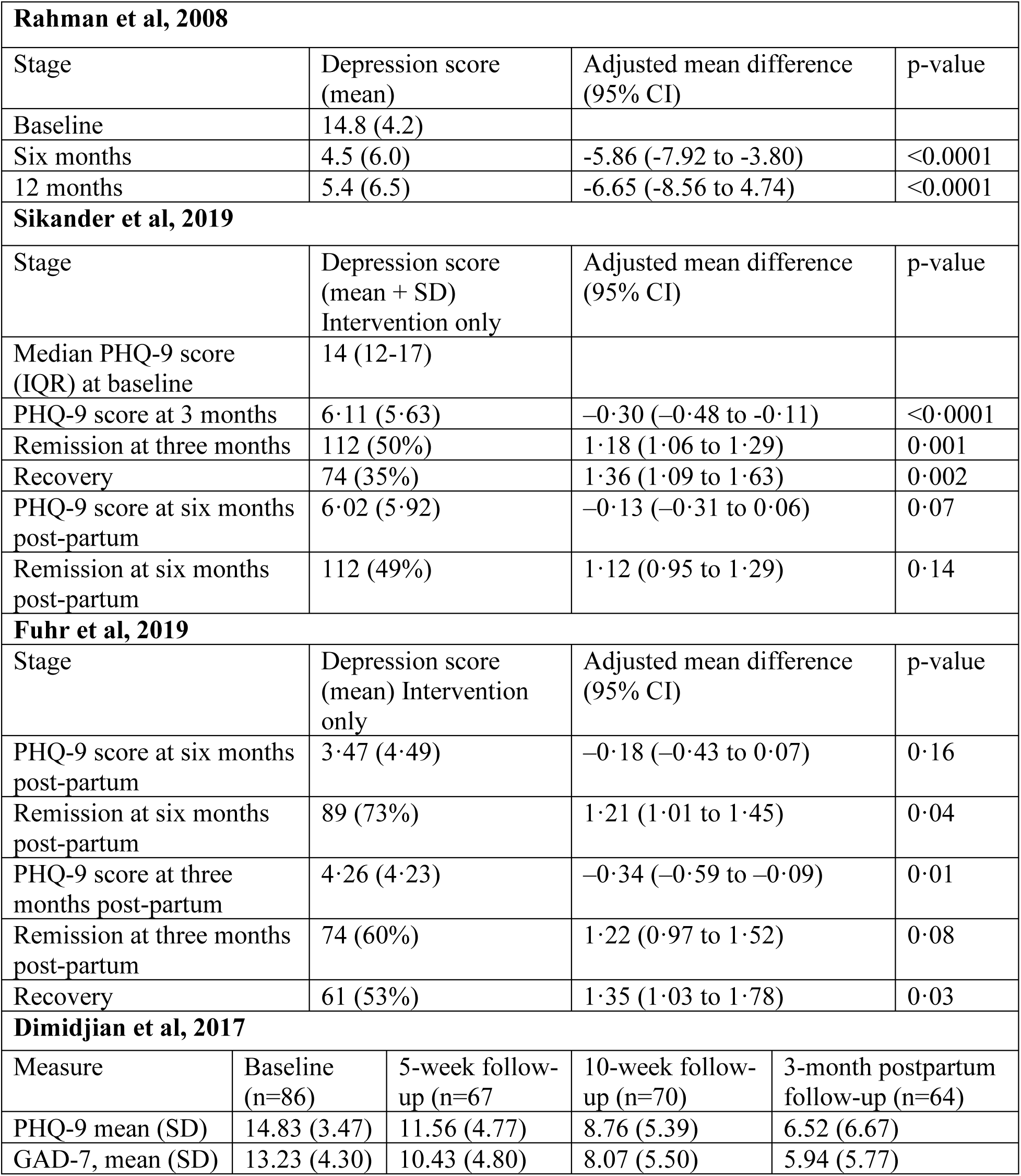
Outcomes the four included studies.

| Rahman et al, 2008 |  |  |  |  |
| --- | --- | --- | --- | --- |
| Stage | Depression score (mean) |  | Adjusted mean difference (95% CI) | p-value |
| Baseline | 14.8 (4.2) |  |  |  |
| Six months | 4.5 (6.0) |  | -5.86 (-7.92 to -3.80) | <0.0001 |
| 12 months | 5.4 (6.5) |  | -6.65 (-8.56 to 4.74) | <0.0001 |
| Sikander et al, 2019 |  |  |  |  |
| Stage | Depression score (mean + SD) Intervention only |  | Adjusted mean difference (95% CI) | p-value |
| Median PHQ-9 score (IQR) at baseline | 14 (12-17) |  |  |  |
| PHQ-9 score at 3 months | 6.11 (5.63) |  | -0.30 (-0.48 to -0.11) | <0.0001 |
| Remission at three months | 112 (50%) |  | 1.18 (1.06 to 1.29) | 0.001 |
| Recovery | 74 (35%) |  | 1.36 (1.09 to 1.63) | 0.002 |
| PHQ-9 score at six months post-partum | 6.02 (5.92) |  | -0.13 (-0.31 to 0.06) | 0.07 |
| Remission at six months post-partum | 112 (49%) |  | 1.12 (0.95 to 1.29) | 0.14 |
| Fuhr et al, 2019 |  |  |  |  |
| Stage | Depression score (mean) Intervention only |  | Adjusted mean difference (95% CI) | p-value |
| PHQ-9 score at six months post-partum | 3.47 (4.49) |  | -0.18 (-0.43 to 0.07) | 0.16 |
| Remission at six months post-partum | 89 (73%) |  | 1.21 (1.01 to 1.45) | 0.04 |
| PHQ-9 score at three months post-partum | 4.26 (4.23) |  | -0.34 (-0.59 to -0.09) | 0.01 |
| Remission at three months post-partum | 74 (60%) |  | 1.22 (0.97 to 1.52) | 0.08 |
| Recovery | 61 (53%) |  | 1.35 (1.03 to 1.78) | 0.03 |
| Dimidjian et al, 2017 |  |  |  |  |
| Measure | Baseline (n=86) | 5-week follow-up (n=67) | 10-week follow-up (n=70) | 3-month postpartum follow-up (n=64) |
| PHQ-9 mean (SD) | 14.83 (3.47) | 11.56 (4.77) | 8.76 (5.39) | 6.52 (6.67) |
| GAD-7, mean (SD) | 13.23 (4.30) | 10.43 (4.80) | 8.07 (5.50) | 5.94 (5.77) |

In the Sikander et al. study, the primary outcomes were symptom severity and remission of depression (defined as a PHQ-9 score of less than 5) at six months post-partum [36]. The secondary outcomes included symptom severity and remission of depression symptoms at three months and recovery (a PHQ-9 score of less than 5) at three and six months [36]. The intervention reduced depression symptom severity (SMD –0·30, 95% CI –0·48 to –0·11; p<0·0001) at three months relative to the control group although reportedly, it did not produce evidence of an effect on symptom severity at six months post-partum. It also did not affect the rate of remission at the same end-point [36] (**Table 2**).

In the Fuhr et al. study, the primary and secondary outcomes were depressive symptom severity, recovery, and remission. Remission and recovery were both defined as a PHQ-9 score of less than five [37]. The intervention reduced depressive symptom severity and improved recovery at three months relative to the control group. There was insufficient evidence of an effect on symptom severity at six months in the intervention group [37]. The prevalence of remission at six months reduced although there was no significant evidence of an intervention effect on remission at three months (p=0·08) [37] (**Table 2**).

The primary outcome of the Dimidjian et al. study was a reduction in depression symptoms. Compared to the control group, participants in the intervention group had their depression symptoms reduced F(1, 152), 4.39; p .04, d, 0.34.d) and there was higher remission (56.3% vs. 30.3%, p .003). The estimated average reduction from baseline across the entire follow-up period was 5.57 (SD 5.03) for BA in the intervention group and 3.98 (SD 5.00) for the control group.

The secondary outcome, anxiety symptom severity, improved for the participants in the intervention group F(1, 152) 6.24; p .014, d 0.41) relative to the control group [35] (**Table 2**).

### Critical Appraisal

We used the revised Cochrane risk-of-bias tool for randomised trials (RoB 2) to assess the risk of bias in the included studies [33]. Of the four included studies, only the Fuhr et al. study scored favourably on the five domains of risk of bias [37]. The common sources of bias were the timing of recruitment of individual participants in relation to the timing of randomisation and measurement of the outcome of interest. In addition, there were concerns about the minimal information provided on the steps taken to ensure fidelity to the intervention (**Figure 2**).

**Figure 2.**
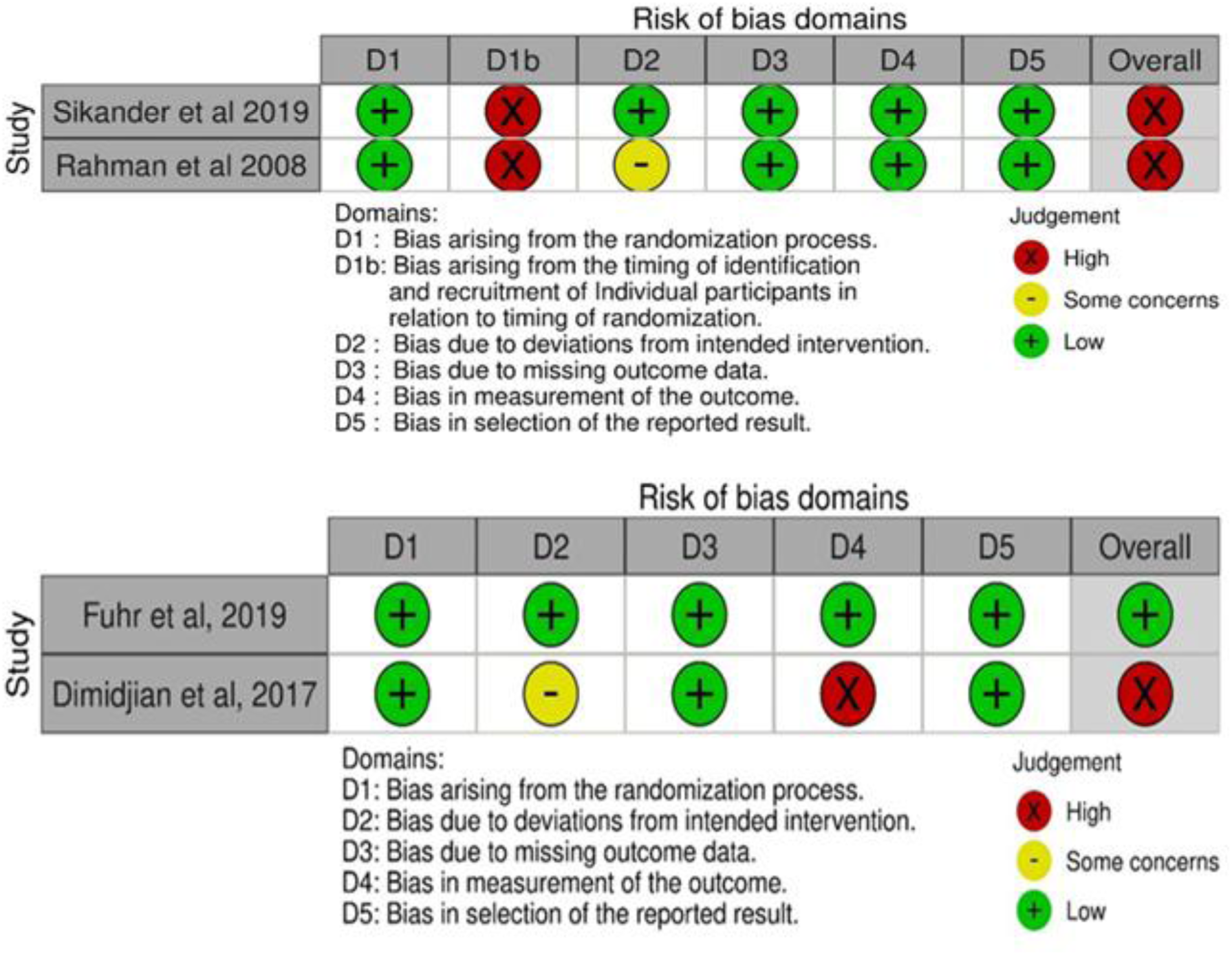
Risk of bias assessment.

### Meta-analysis and heterogeneity test

We performed a meta-analysis to understand the effects of behavioural activation versus TAU on treating prenatal depression. The overall results showed that compared to TAU, behavioural activation resulted in a modest pooled effect size on depressive symptoms in women during the prenatal period [Standardised mean difference of −0.33 (95% CI, −0.57 to −0.10)] (**Figure 3**).

**Figure 3.**
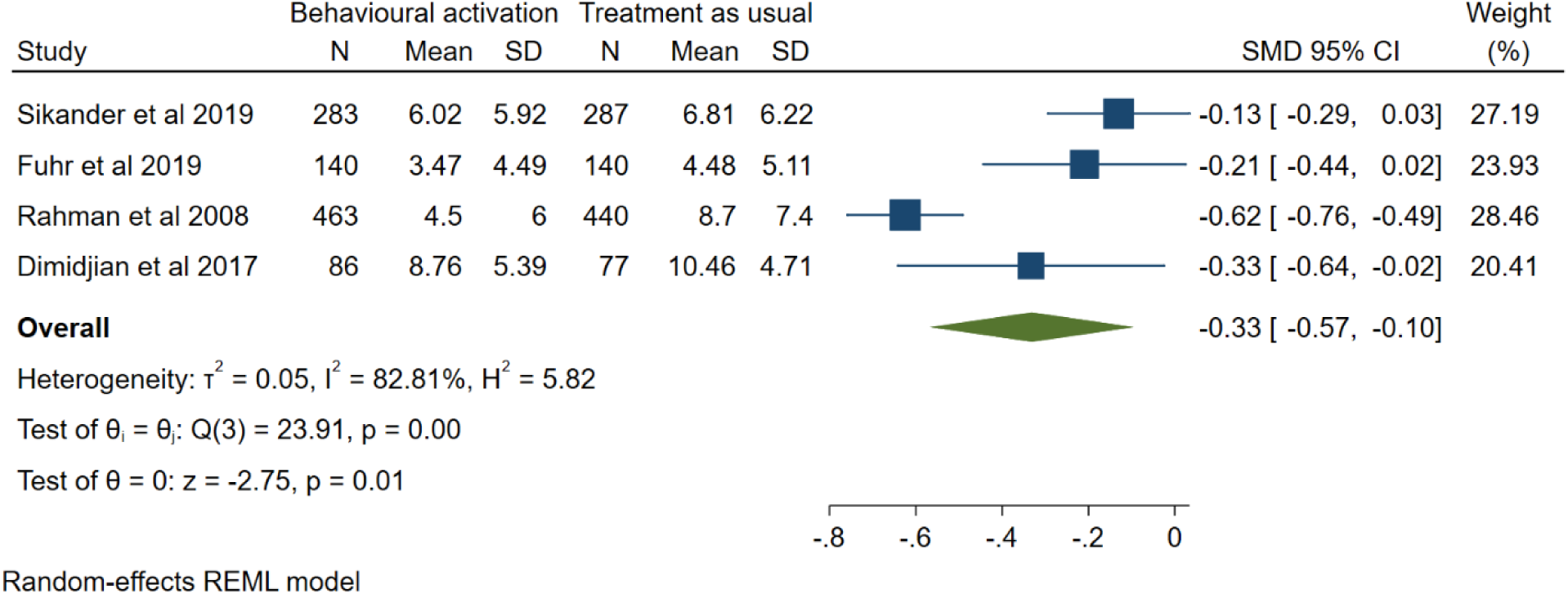
Meta-analysis and heterogeneity test showing the effect of BA on prenatal depression.

The meta-analysis included an assessment of heterogeneity. The results of the heterogeneity tests showed substantial disparity among the included studies.

### Subgroup analysis

We performed subgroup analyses to understand the possible source of heterogeneity linked with the pooled estimate of the effects of behavioural activation on depressive symptoms based on study design, the type of outcome measures, and how the treatment is defined. The forest plots from these subgroup analyses are presented in **Figures 4a-4c**.

**Figure 4a.**
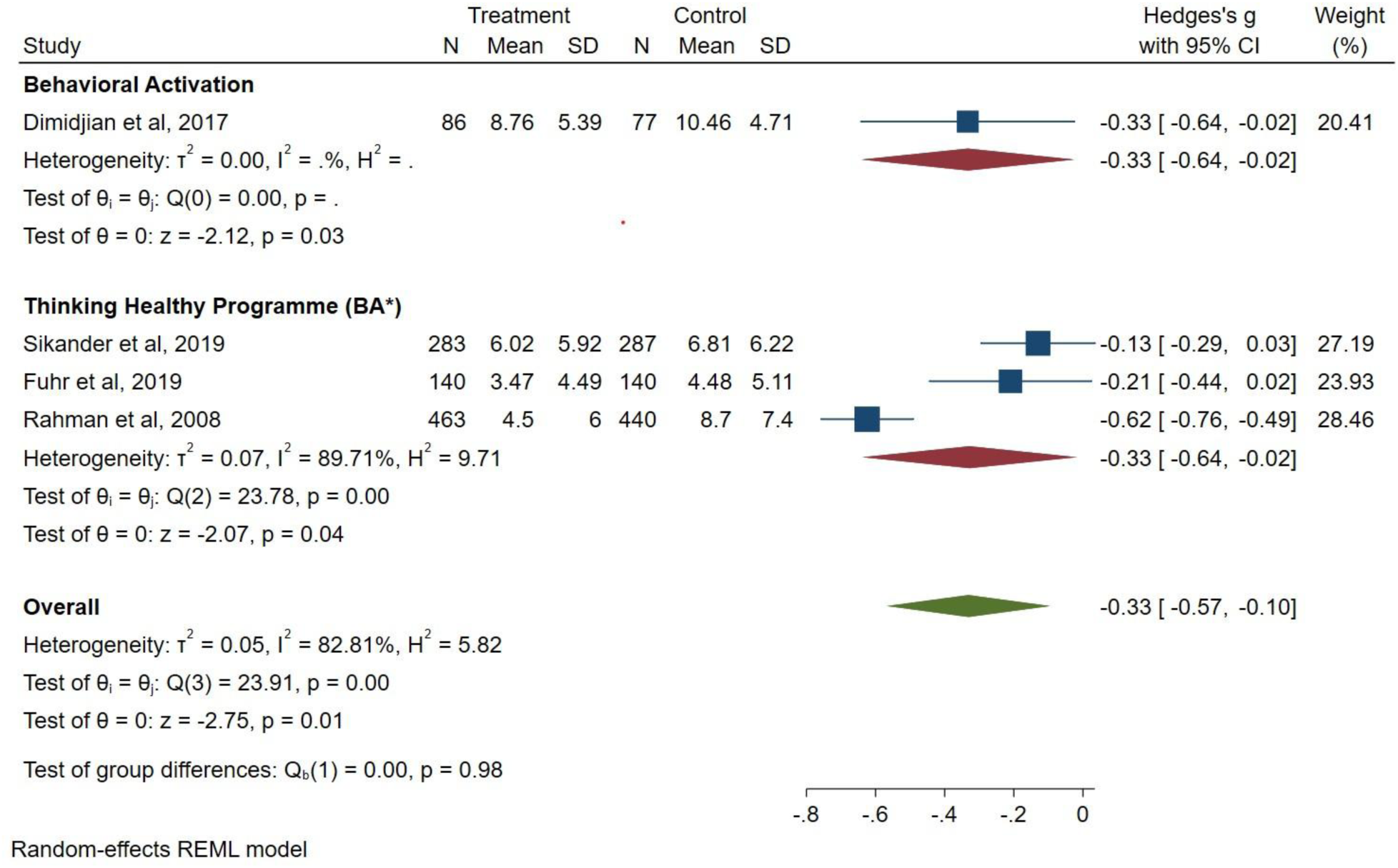
Sub-group analysis by study design.

**Figure 4b.**
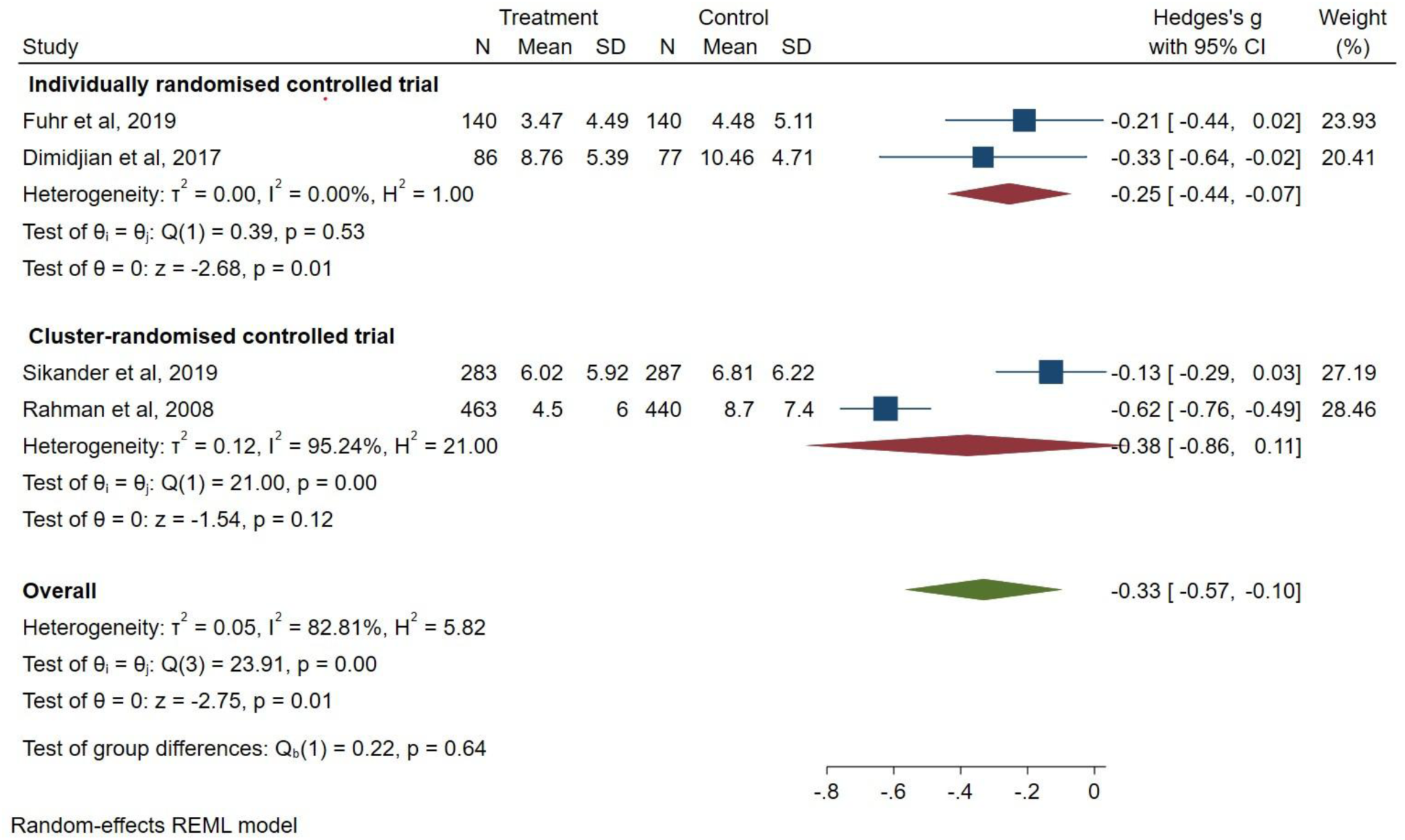
Sub-group analysis by intervention type.

**Figure 4c.**
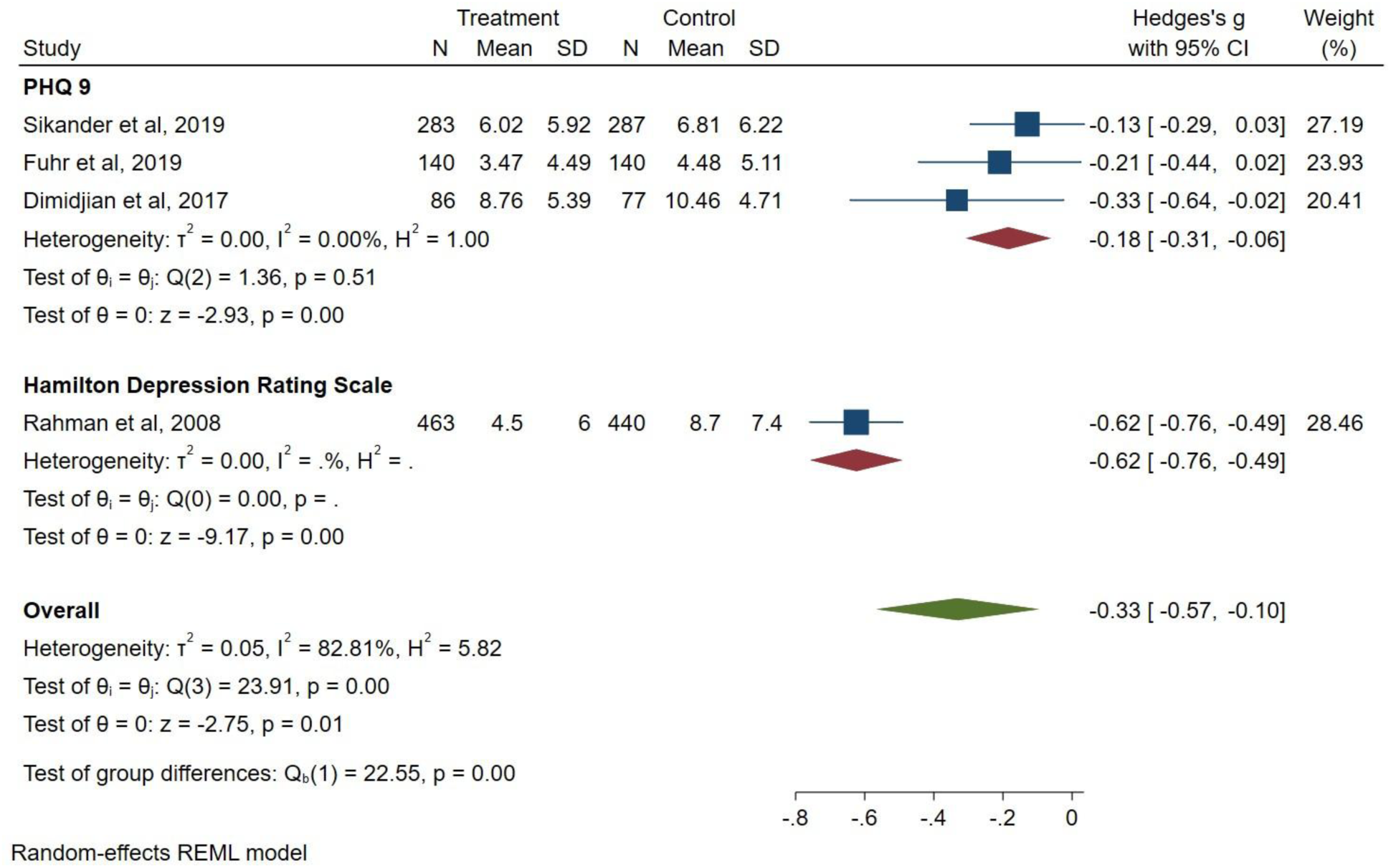
Sub-group analysis by outcome measure.

### Reporting of adverse events and harms

Two of the included studies reported adverse events such as the death of the child, hospital admissions of the child, and experience of physical violence [36, 37]. None of the included studies reported any harms caused to participants due to participation in the treatment trials.

### Deviations from systematic review protocol

Nothing to report.

### Potential biases in the review process

Our search strategy included global databases listing publications in English. In addition, we included studies in which behavioural activation was a component and not the core element of the intervention. This means interventions in other languages or relevant to clinical practice, such as collaborative care without substantial behavioural activation, may have been excluded.

## Discussion

In this systematic review, we aimed to summarise the evidence about the efficacy of behavioural activation compared with usual care in treating prenatal depression. Behavioural activation is already recommended for the treatment of depression in adults [16] due to its advantages over other psychological therapies [19]. We included results from four randomised controlled trials that recruited 972 participants from India, Pakistan, and the USA [34–37].

In all four included studies, behavioural activation was effective in improving depressive symptoms during the prenatal period. This evidence suggests that there may be potential for using behavioural activation in treating depression in women during the prenatal period despite weaknesses due to study biases, small samples, and a paucity of evidence. The present evidence suggests that behavioural activation may not only be effective in treating depression but also helpful in improving remission from the psychological condition. In one study, behavioural activation had a significant effect on anxiety symptoms [35].

There is a growing focus on scaling up global and public health interventions for optimal reach and outcomes [49, 50]. The argument has been put forward for public health interventions to incorporate scale in their design and implementation [51]. Yet scaling-up of interventions for managing depression in adults often remains challenged by the lack of an appropriately qualified and skilled specialist health workforce due to widespread and persistent shortage and maldistribution [52, 53]. We define scaling-up as a deliberate effort to increase the impact of health service interventions that have been successfully tested in pilot projects so that they will benefit more people on a lasting basis [51, 54]. We argue that interventions may be of limited value unless they can be demonstrated to generate wider benefits and impact. Specifically, this is the gap that behavioural activation may be capable of addressing in treating depression in women prenatally due to its capacity for scale-up. We speculate that this finding may have relevance for resource-poor settings and environments with high prevalence and incidence of prenatal depression, especially low- and middle-income countries. Evidence of sustainability of the behavioural activation interventions for depressed women was inconclusive.

Of the four included studies, only one study had a low risk of bias. Biases were observed concerning the timing of recruitment of individual participants in relation to the timing of randomisation and measurement of the outcome of interest. Future studies may want to pay attention to the strengthening of methodological designs and processes.

We recognise that the evidence from this review may not be sufficient to conclude the efficacy of behavioural activation in treating depression in women during the prenatal period. Further research is required to grow robust evidence on the use of behavioural activation as a viable alternative to other treatments for prenatal depression in women.

### Limitations of this review

The small number of included studies would suggest that caution should be exercised in interpretation [55]. Furthermore, the results of this review may not be generalisable due to the small sample size. Publication bias may have ensued from the inclusion of papers published in English only because we wanted to avoid challenges with translation. The review also excluded grey literature.

## Conclusions

Depression in women during the prenatal period can induce severe consequences for the mother, the child, and the wider family network. Our systematic review shows that behavioural activation may be an efficacious treatment for prenatal depression. The methodological biases identified in this review suggest a need for caution in interpreting the results. Further research is required to build up robust evidence about the efficacy and sustainability of behavioural activation interventions for prenatal depression. Based on the evidence from the four studies included in this review, opportunities exist for different types of non-specialist mental health workers to be prepared to deliver behavioural activation to women during the prenatal period.

## Abbreviations

BA: Behavioural activation
BADS-SF: Behavioural Activation for Depression Scale-Short Form
CBT: Cognitive Behavioural therapy
CI: Confidence interval
DSM-IV: Diagnostic and Statistical Manual of Mental Disorders
GAD-7: Generalised Anxiety Disorder 7-item Questionnaire
PHQ-9: Patient Health Questionnaire-9
PICO: Population-Intervention-Comparison-Outcome
PRISMA: Preferred Reporting Items for Systematic Reviews and Meta-Analyses
SMD: Standardised mean difference
TAU: Treatment as usual
THPP: Thinking Healthy Programme
WHO: World Health Organization

## Data Availability

All data produced in the present work are contained in the manuscript.

## Acknowledgements

The authors acknowledge the assistance of academic librarian Lorien Delaney of the University of South Australia. Lorien assisted with the preparation of the search strategy and conducting database searches.

## Author contribution

KM, EY, MJ, and MS conceived the project. MJ, SW, EY, SO, and KM performed searches of relevant data sources, screened identified publications for inclusion, and extracted the data reported in this manuscript. EY and MJ completed the quality appraisal of the included studies. EY performed the meta-analysis. KM and EY prepared the initial version of the manuscript. MJ, MS, DB, JF, SW, SO, KM, and EY edited various drafts of the manuscript. All authors reviewed and approved this manuscript. EY is the guarantor.

## Data availability statement

Materials supporting the findings reported in this manuscript are available from the corresponding author upon reasonable request.

## Declarations

### Ethical considerations

Not applicable

### Consent for publication

Not applicable.

## Supplementary material

### Appendix 1. MEDLINE Search Strategy

Pregnancy/

Peripartum Period/

Postpartum Period/

Puerperal Disorders/

(pregnan* or ante natal or antenatal or post partum or postpartum or ante partum or antepartum or post natal or postnatal or peri partum or peripartum or peri natal or perinatal or intra partum or intrapartum or puerp* or after birth or pre natal or prenatal).ti,ab,kf.

or/1-5

depression/

mood disorders/

depressive disorder/

depressive disorder, major/

depressive disorder, treatment-resistant/

dysthymic disorder/

cyclothymic disorder/

(depress* or dysthymi* or cyclothymi* or low mood or mood disorder* or affective disorder*).ti,ab,kf.

or/7-14

6 and 15

Depression, Postpartum/

16 or 17

Behavior Therapy/

Motivational Interviewing/

(behavio* activat* or BATD).ti,ab,kf.

(behavio* adj3 (reinforce* or re-inforce*)).ti,ab,kf.

(behavio* adj2 (contracting or modif*)).ti,ab,kf.

reinforc*.ti,kf. or ((positive adj1 reinforc*) or (reinforc* adj3 (environment* or experience*))).ti,ab,kf.

((behavio* adj3 motivat*) or motivational interviewing).ti,ab,kf. (activit* adj2 schedul*).ti,ab,kf.

((pleas* or enjoyable or rewarding) adj3 (activit* or event or events)).ti,ab,kf. ((operant or instrumental) adj (conditioning or learning)).ti,ab,kf.

(positive interaction* or avoida* coping or environmental contingenc* or contingency management).ti,ab,kf.

functional analysis.ti,ab,kf.

behavio*.mp. and (self adj (evaluat* or monitor*)).ti,ab,kf.

(behavio* adj (counsel* or intervention* or train* or treatment* or therap* or psychotherap*)).ti,ab,kf.

(mood adj3 monitor*).ti,ab,kf.

or/19-33

18 and 34

randomized controlled trial.pt.

controlled clinical trial.pt.

randomized.ab.

placebo.ab.

drug therapy.fs.

randomly.ab.

trial.ab.

groups.ab.

or/36-43

exp animals/ not humans.sh.

44 not 45

35 and 46

### Appendix 2. A detailed summary of the included studies

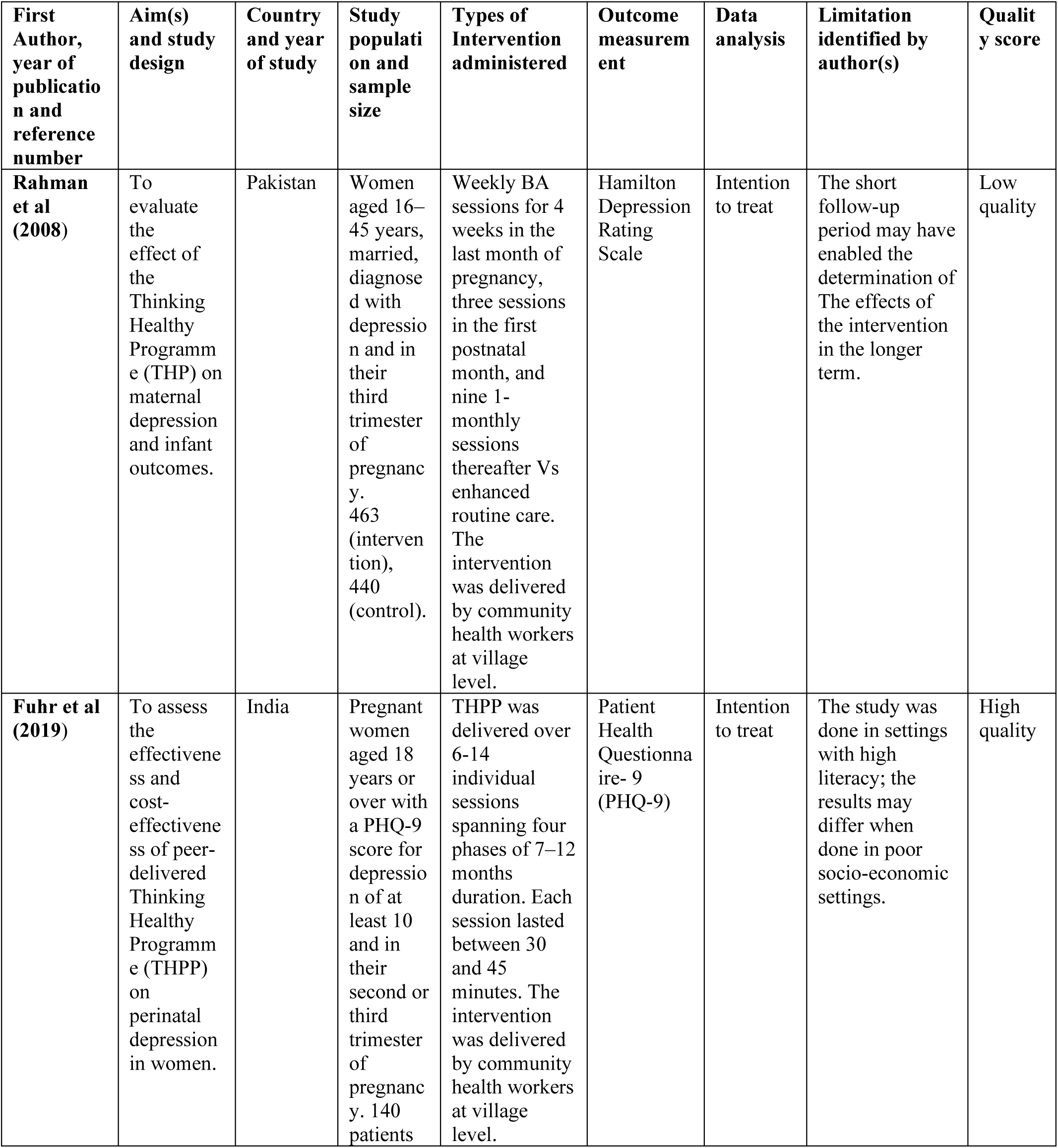
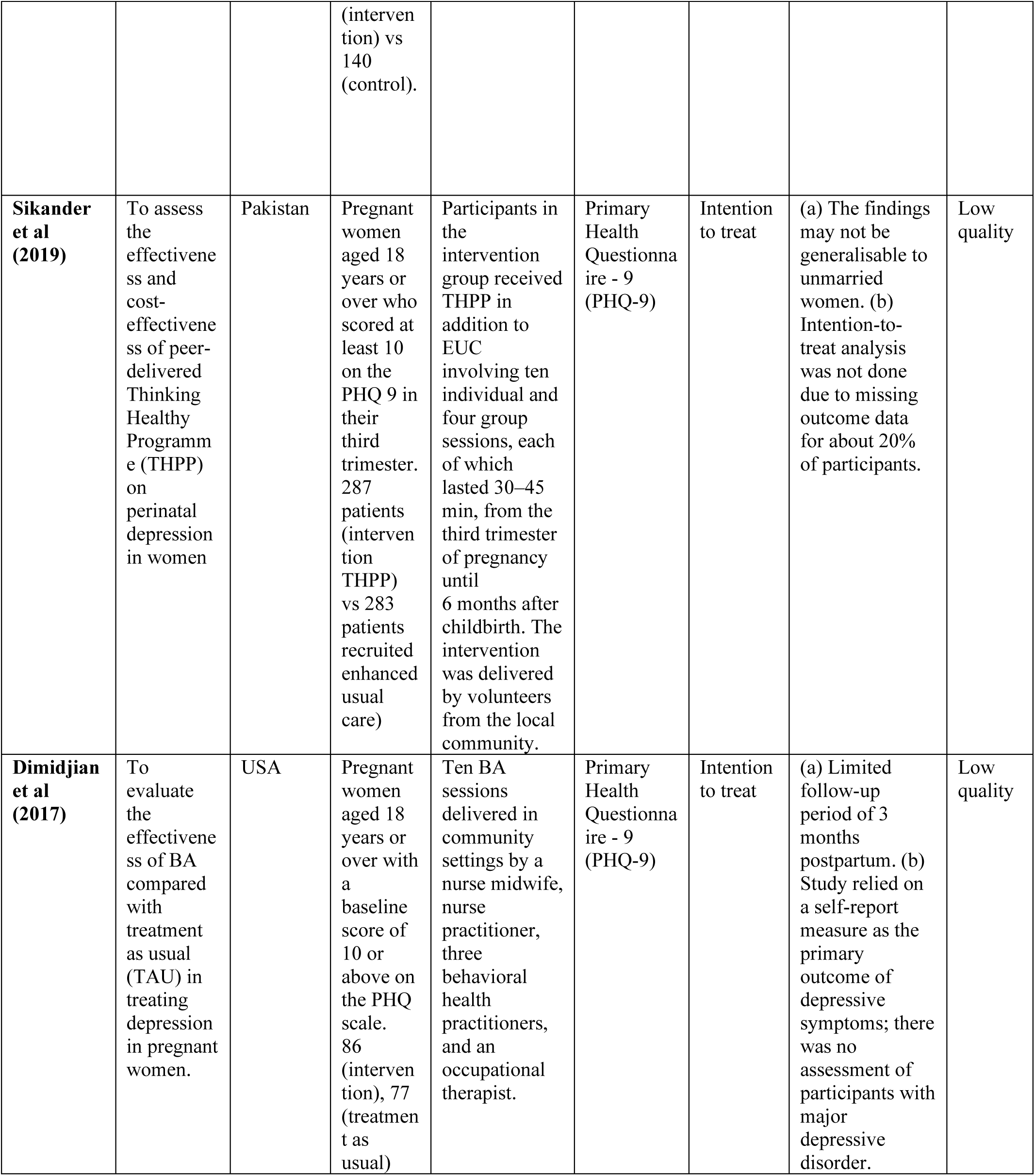

| First Author, year of publication and reference number | Aim(s) and study design | Country and year of study | Study population and sample size | Types of Intervention administered | Outcome measurement | Data analysis | Limitation identified by author(s) | Quality score |
| --- | --- | --- | --- | --- | --- | --- | --- | --- |
| **Rahman et al (2008)** | To evaluate the effect of the Thinking Healthy Programme (THP) on maternal depression and infant outcomes. | Pakistan | Women aged 16–45 years, married, diagnosed with depression and in their third trimester of pregnancy. 463 (intervention), 440 (control). | Weekly BA sessions for 4 weeks in the last month of pregnancy, three sessions in the first postnatal month, and nine 1-monthly sessions thereafter Vs enhanced routine care. The intervention was delivered by community health workers at village level. | Hamilton Depression Rating Scale | Intention to treat | The short follow-up period may have enabled the determination of The effects of the intervention in the longer term. | Low quality |
| **Fuhr et al (2019)** | To assess the effectiveness and cost-effectiveness of peer-delivered Thinking Healthy Programme (THPP) on perinatal depression in women. | India | Pregnant women aged 18 years or over with a PHQ-9 score for depression of at least 10 and in their second or third trimester of pregnancy. 140 patients (intervention) vs 140 (control). | THPP was delivered over 6-14 individual sessions spanning four phases of 7–12 months duration. Each session lasted between 30 and 45 minutes. The intervention was delivered by community health workers at village level. | Patient Health Questionnaire- 9 (PHQ-9) | Intention to treat | The study was done in settings with high literacy; the results may differ when done in poor socio-economic settings. | High quality |
| **Sikander et al (2019)** | To assess the effectiveness and cost-effectiveness of peer-delivered Thinking Healthy Programme (THPP) on perinatal depression in women | Pakistan | Pregnant women aged 18 years or over who scored at least 10 on the PHQ 9 in their third trimester. 287 patients (intervention THPP) vs 283 patients recruited enhanced usual care) | Participants in the intervention group received THPP in addition to EUC involving ten individual and four group sessions, each of which lasted 30–45 min, from the third trimester of pregnancy until 6 months after childbirth. The intervention was delivered by volunteers from the local community. | Primary Health Questionnaire - 9 (PHQ-9) | Intention to treat | (a) The findings may not be generalisable to unmarried women. (b) Intention-to-treat analysis was not done due to missing outcome data for about 20% of participants. | Low quality |
| **Dimidjian et al (2017)** | To evaluate the effectiveness of BA compared with treatment as usual (TAU) in treating depression in pregnant women. | USA | Pregnant women aged 18 years or over with a baseline score of 10 or above on the PHQ scale. 86 (intervention), 77 (treatment as usual) | Ten BA sessions delivered in community settings by a nurse midwife, nurse practitioner, three behavioral health practitioners, and an occupational therapist. | Primary Health Questionnaire - 9 (PHQ-9) | Intention to treat | (a) Limited follow-up period of 3 months postpartum. (b) Study relied on a self-report measure as the primary outcome of depressive symptoms; there was no assessment of participants with major depressive disorder. | Low quality |

---

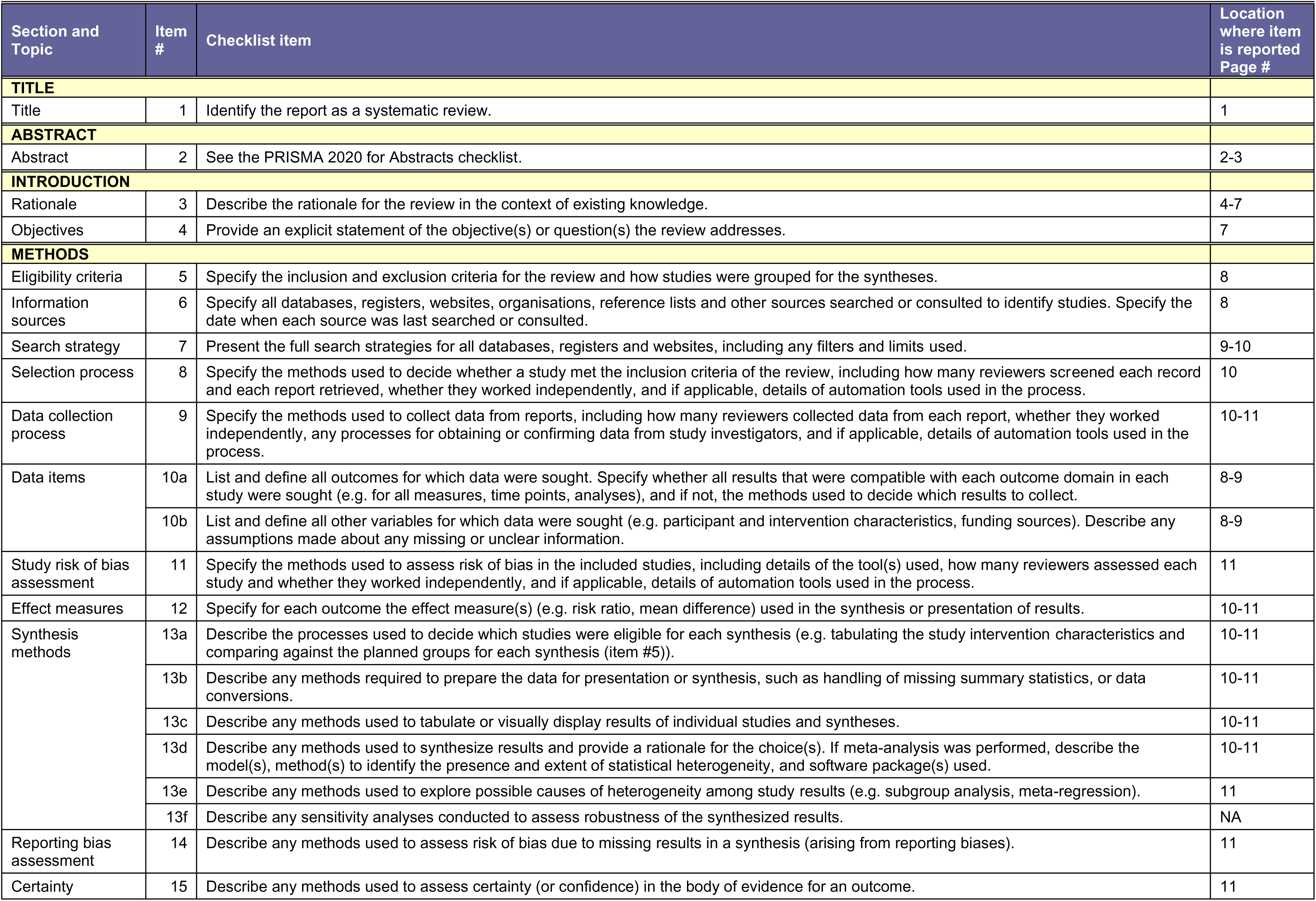
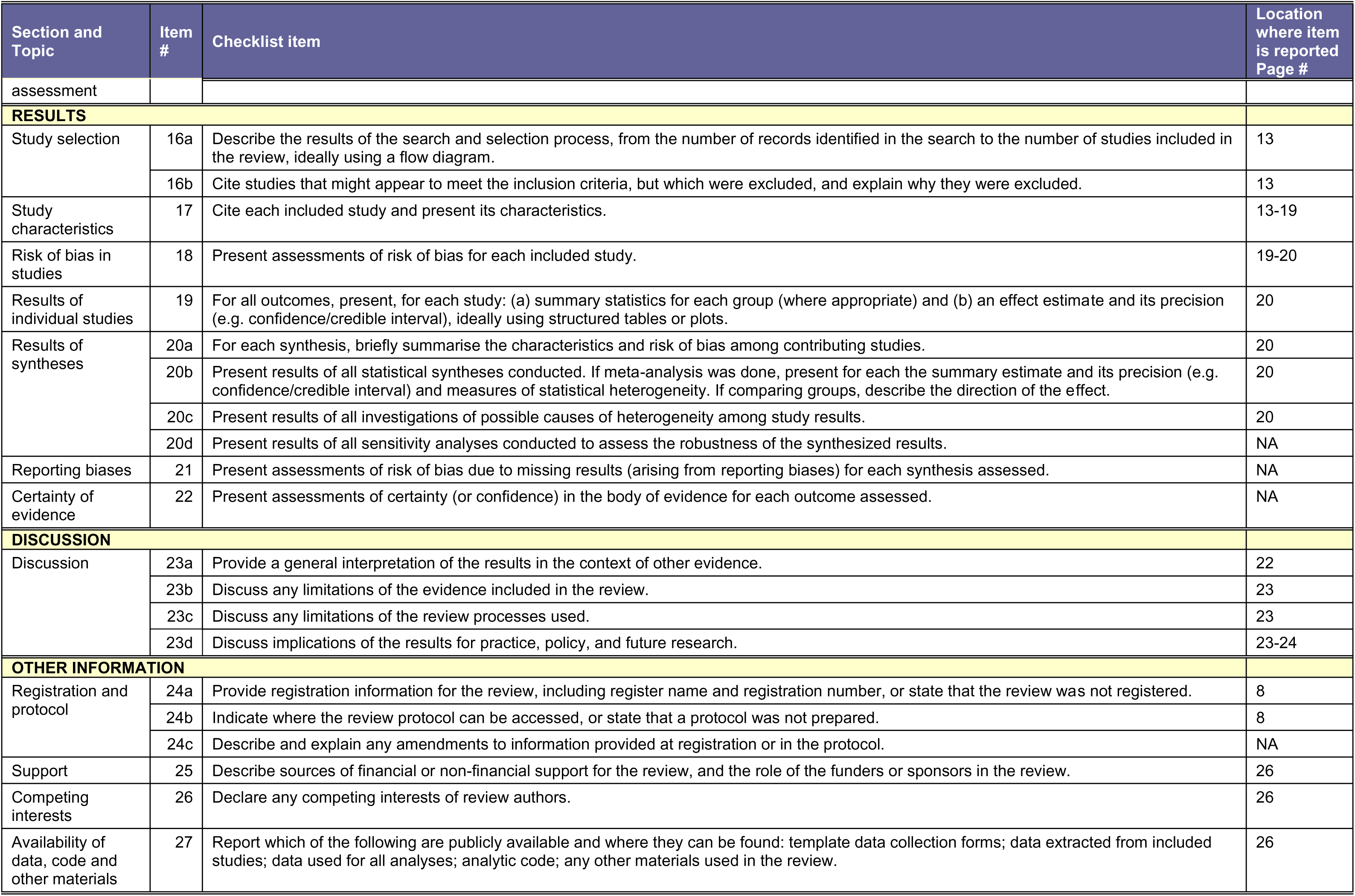
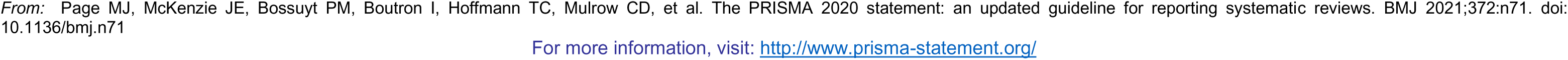

